# A Comparative Study Between Nasopharyngeal/Oropharyngeal, Faecal and Saliva Viral Shedding In Ghanaian COVID-19 Patients attending Komfo Anokye Teaching Hospital (KATH), Kumasi from October – December, 2020

**DOI:** 10.1101/2021.09.04.21262932

**Authors:** Ernest Badu-Boatng, Lydia Sarponmaa Asante, Albert Dompreh, Laud Anthony Basing W., Kwabena Adjei Asante, Sylvia Karikari, Albert Adubofour, Chris Oppong, Faustina Acheampong

## Abstract

**Background:** Diagnostic testing for the current SARS-CoV-2 infections involves the collection and testing of invasive pharyngeal specimens by qualified Health workers. Though fully clad in personal protective equipment, the concern is that sampling in close proximity to the patient poses as a major health hazard. The present study sought to verify if saliva or faeces could become a possible surrogate for pharyngeal samples for SARS-CoV 2 testing in suspected Ghanaian COVID-19 patients.

**Objectives:** To ascertain if there is SARS-CoV 2 viral shedding in the saliva and faecal samples of Ghanaian COVID-19 patients, their sensitivity and specificity as compared to pharyngeal samples.

**Method:** Fifty (50) recruited COVID-19 patients who have been confirmed via RT-PCR using their nasopharyngeal/oropharyngeal samples and twenty (20) SARS-CoV 2 negative suspected patients each provided some faecal and saliva sample for RT-PCR analysis for SARS-CoV 2.

**Results:** Forty-three (43) out of the fifty (50) COVID-19 patients recruited representing 86% tested positive for SARS-CoV 2 via their saliva sample whiles all their faecal samples tested positive for SARS-CoV 2 representing 100%. The sensitivity of saliva samples was 86% whiles the specificity was 100% but the sensitivity and specificity of the faecal samples were all 100%.

**Conclusion:** There is indeed viral shedding of SARS-CoV 2 in the saliva and faeces of Ghanaian COVID-19 patients just like their counterparts in other parts of the world. Saliva and faeces could possibly become an alternative sample to the current in place of the invasive pharyngeal samples for SARS-CoV 2 testing in resource limited settings. Further research to explore this possibility at different testing sites with larger sample size is recommended.

## INTRODUCTION

### 1.1 Background

The Novel Corona Virus (SARS-CoV 2) was confirmed by Health Authorities in the Chinese city of Wuhan (Al-Qahtani, 2020). This is within the Hubei province in The People’s Republic of China (Zhu et al., 2020). The confirmation came in late December, 2019 after a series of patients reported with pneumonia of unknown origin (Zhu et al., 2020). Initial reported cases were linked to a local seafood and wild animal market which was suggestive of a zoonotic transmission (Walls et al., 2020). It soon became evident that this unknown virus (SARS-CoV 2) was spreading from person to person (Yuen et al., 2020). Health workers treating the identified cases equally developed symptoms (Zhu et al., 2020). It got sequenced and isolated in January, 2020 (*Whole Genome of Novel Coronavirus, 2019-NCoV, Sequenced -- ScienceDaily*, 2020) and is associated with the current ongoing global pandemic called Corona Virus Disease-2019 (Lo et al., 2020).

Coronaviruses are a large family of enveloped, non-segmented, positive-sense single-stranded Ribonucleic Acid (RNA) viruses (Pal et al., 2020). In the 1960s, humans were getting infected with corona viruses (Walsh et al., 2013) causing common cold and respiratory illnesses especially during winter months in temperate climates (Walsh et al., 2013). Individuals with COVID-19 usually develop signs and symptoms, such as mild respiratory illness and persistent fever in about 5-6 days after infection on the average but ranges between 1-14 days (De Oliveira Lima, 2020). Three coronaviruses have crossed the species barrier to cause deadly pneumonia in humans since the beginning of the 21^st^ century (Walls et al., 2020).

These are Severe Acute Respiratory Syndrome Corona Virus (SARS-CoV), Middle East Respiratory Syndrome Corona Virus (MERS-CoV) and Severe Acute Respiratory Syndrome Corona Virus-2 [SARS-CoV 2] (Walls et al., 2020).

SARS-CoV emerged from the Guangdong Province of China in 2002 spreading to five (5) continents through air-travel routes (Jiang et al., 2005). It infected 8,450 people and killed 810 of them worldwide in 33 countries (Jiang et al., 2005). MERS-CoV also emerged in 2012 in the Arabian Peninsula, spreading to 27 Countries, infected 2,494 people with 858 deaths (Walls et al., 2020). SARS-CoV 2 enters the host cells using the Angiotensin-Converting Enzyme 2 [ACE2] (Hoffmann et al., 2020). The spike-glycoprotein of SARS-CoV 2 which is a transmembrane spike forms homotrimers (Astuti & Ysrafil, 2020). This protrudes from the viral surface to act as the functional receptor binding to the ACE2 for entry into the host cells (Astuti & Ysrafil, 2020). This is because of their high affinity for each other (Astuti & Ysrafil, 2020).

**Figure 1:**
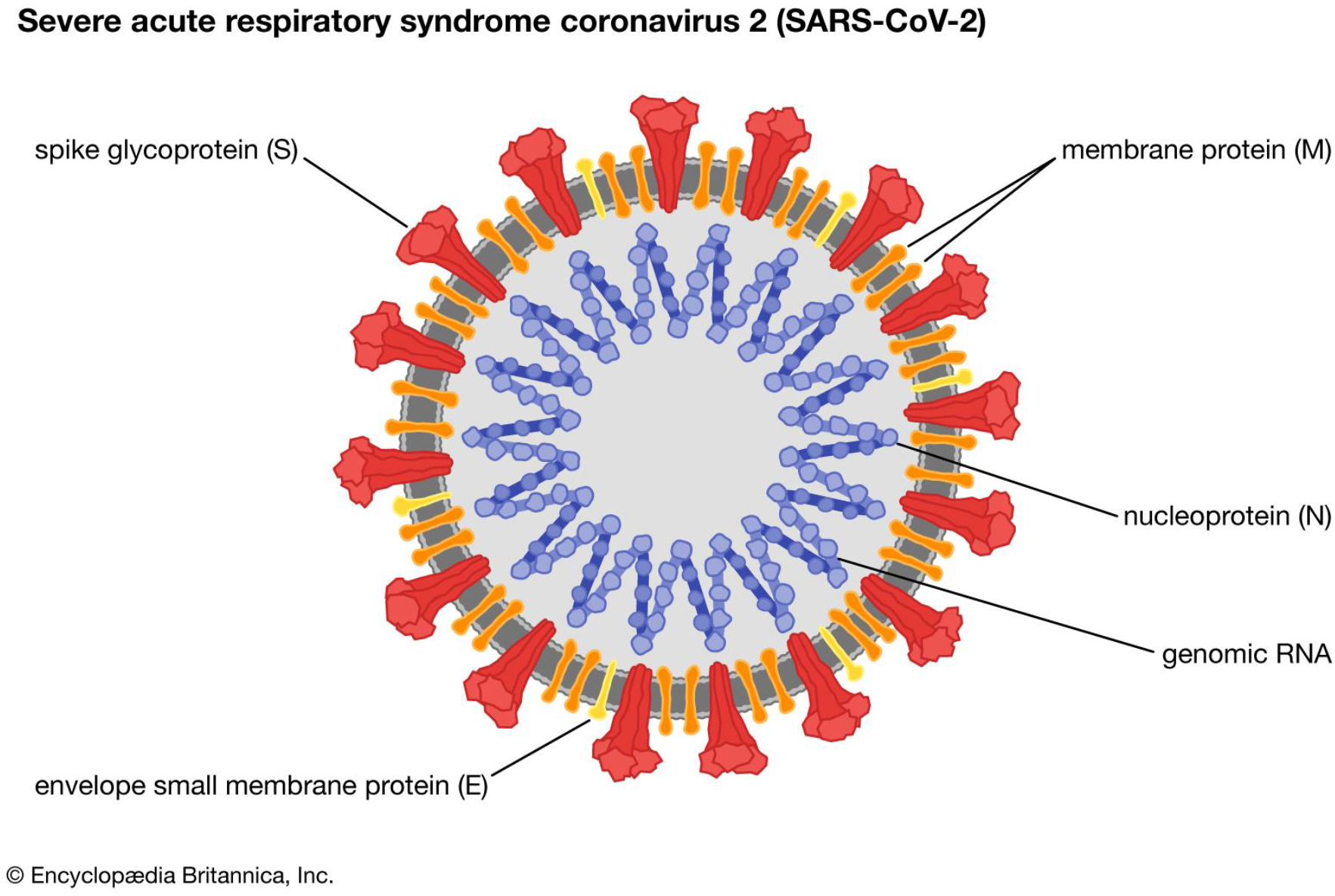
Structure of SARS-CoV 2 (Coronavirus | Definition, Features, & Examples | Britannica, 2020)

The spike-glycoprotein is surface exposed and plays this vital facilitative role of the viral entry into susceptible host cells (Du et al., 2009). It has therefore become the main target of neutralizing antibodies upon infection (Du et al., 2009) as well as the focus of Therapeutic and Vaccine Designers (Du et al., 2009). The ACE2 messenger RNA is highly expressed and stabilized by the presence of a neutral amino acid transporter in the gastrointestinal system (Perlot, 2013). Possible activation of SARS-CoV 2 virus by gene expression of furin in salivary glands is a noteworthy finding (Ibrahim Warsi, 2021). Furin is typically expressed by salivary glands and its components are responsible for the regulation of different specific proteins while the gene itself cleave different viral toxins including coronaviruses (Ibrahim Warsi, 2021). Infection normally takes place from person to person through droplets from coughs, sneezes or from contaminated hands and surfaces [fomites] (Jayaweera et al., 2020).

**Figure 2:**
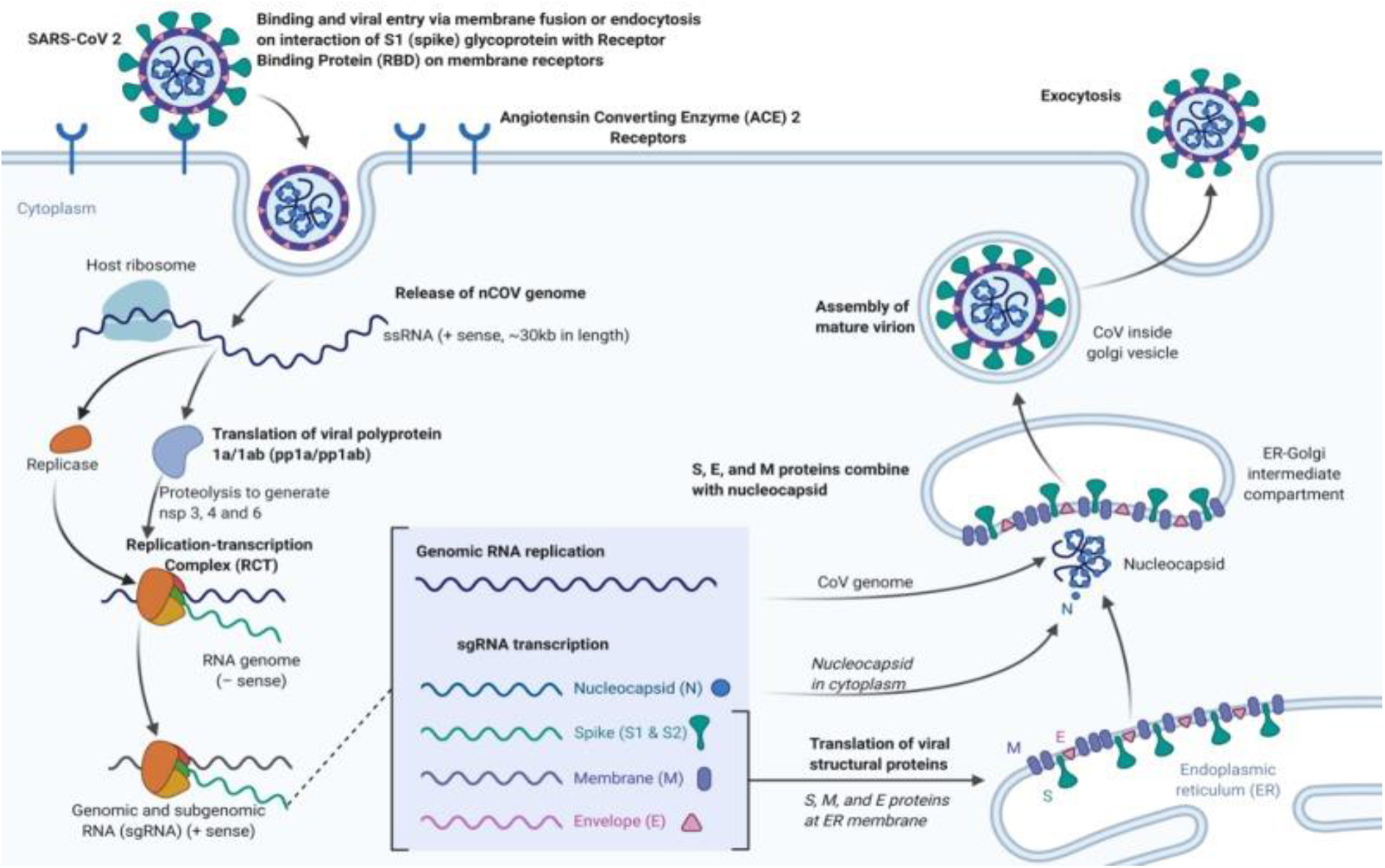
Replication of SARS-CoV 2 in the Human host (Transmission, Pathogenesis, Replication of SARS-CoV-2 (COVID-19), 2020)

The WHO reported global COVID-19 confirmed cases of 9,473,214 with 484,249 deaths as at 26^th^ June, 2020 at 10:00 CEST (*WHO Coronavirus Disease (COVID-19) Dashboard | WHO Coronavirus Disease (COVID-19) Dashboard*, 2020). Out of this global figure of infected patients reported by the WHO, the Americas lead the global confirmed cases of COVID-19 at that very time with 4,709,924 and 233,628 deaths, followed by Europe with 2,619,753 confirmed cases and 195,535 deaths, South-East Asia with 686,192 confirmed cases and 19,651 deaths. Eastern Mediterranean had 987,534 confirmed cases with 22,464 deaths (*WHO Coronavirus Disease (COVID-19) Dashboard | WHO Coronavirus Disease (COVID-19) Dashboard*, 2020).

Africa had 258,752 confirmed cases with 5,564 deaths as at the time followed closely by Western Pacific with the least number of confirmed cases being 210,315 with 7,394 deaths (*WHO Coronavirus Disease (COVID-19) Dashboard | WHO Coronavirus Disease (COVID-19) Dashboard*, 2020). From the WHO global data at the time, a lot of the cases being reported were coming from community transmission or spread (*WHO Coronavirus Disease (COVID-19) Dashboard | WHO Coronavirus Disease (COVID-19) Dashboard*, 2020). In sub-Saharan Africa including Ghana, South Africa, Ethiopia, Nigeria, Kenya, Cote d’Ivoire, Burkina Faso, Benin, Gambia, Guinea, Togo, Niger, Mali, Sierra Leone and Liberia just to mention a few, a total of over 188,300 confirmed cases has been recorded as at 26^th^ June, 2020 with over 3,591 deaths unfortunately (*WHO Coronavirus Disease (COVID-19) Dashboard | WHO Coronavirus Disease (COVID-19) Dashboard*, 2020).

Ghana as at 4^th^ August, 2020 had reported a total of 40,533 confirmed COVID-19 cases since the first two cases were announced on 12^th^ March, 2020 (*COVID-19 Updates | Ghana*, 2020) with over 37,702 Discharges/Recoveries, 206 Deaths and 2,625 Active cases (*COVID-19 Updates | Ghana*, 2020).

The regional case count at the time put Greater Accra Region on top of the chart with 20,413 confirmed cases (*COVID-19 Updates | Ghana*, 2020) followed by Ashanti Region with 10,097 confirmed cases (*COVID-19 Updates | Ghana*, 2020). Western Region had recorded 2,796 confirmed cases followed by Eastern Region with 1,846 confirmed cases (*COVID-19 Updates | Ghana*, 2020). Central Region had 1,669 confirmed cases with Bono East Region also having 636 confirmed cases (*COVID-19 Updates | Ghana*, 2020). Volta Region had recorded 617 confirmed cases with Western North Region recording 557 confirmed cases by 4^th^ August, 2020 (*COVID-19 Updates | Ghana*, 2020).

Northern Region has recorded 454 confirmed cases with Bono Region also having recording 439 confirmed cases (*COVID-19 Updates | Ghana*, 2020). Ahafo Region has recorded 364 confirmed cases with Upper East Region recording 282 confirmed cases (*COVID-19 Updates | Ghana*, 2020). Oti Region has recorded 204 confirmed cases with Upper West Region also recording 88 confirmed cases (*COVID-19 Updates | Ghana*, 2020). Savannah Region has recorded 62 confirmed cases with North East Region recording 9 confirmed cases (*COVID-19 Updates | Ghana*, 2020). The gender distribution of all these confirmed cases is greater in Males (60%) than Females (40%) (*COVID-19 Updates | Ghana*, 2020).

Coronavirus is already impacting global markets, with economic impacts felt beyond China and other effected countries (*What Impact Is the Coronavirus Having on the Global Economy? | World Economic Forum*, 2020). As the human toll mounts, so does the economic damage (*What Impact Is the Coronavirus Having on the Global Economy? | World Economic Forum*, 2020). Most governments around the world temporarily closed educational institutions in an attempt to contain the spread of the virus (*School Closures Caused by Coronavirus (Covid-19)*, 2020).

These nationwide closures impacted hundreds of millions of students (*School Closures Caused by Coronavirus (Covid-19)*, 2020). The potent combination of trip cancellations and country-specific restrictions on international flights had a staggering impact on the $880 billion global airline industry (*Has Coronavirus Stopped All Flights? | World Economic Forum*, 2020). Around 90% of commercial passenger flights are grounded (Devi, 2020).

More than 130 countries introduced some form of travel restriction since this outbreak began, including screening, quarantine, and travel ban from high-risk areas (Devi, 2020).

In Sub-Saharan Africa, economic growth has been significantly impacted by the ongoing coronavirus outbreak (*COVID-19 (Coronavirus) Drives Sub-Saharan Africa Toward First Recession in 25 Years*, 2020) and is forecasted to fall sharply from 2.4% in 2019 to −2.1 to −5.1% in 2020 (*COVID-19 (Coronavirus) Drives Sub-Saharan Africa Toward First Recession in 25 Years*, 2020), the first recession in the region over the past 25 years, according to the latest Africa’s Pulse, the World Bank’s twice-yearly economic update for the region (*COVID-19 (Coronavirus) Drives Sub-Saharan Africa Toward First Recession in 25 Years*, 2020).

The analysis showed that COVID-19 will cost the region between $37 billion and $79 billion in output losses for 2020 due to a combination of effects (*COVID-19 (Coronavirus) Drives Sub-Saharan Africa Toward First Recession in 25 Years*, 2020).

Ghana has not been left out of the many, if not all stock markets that have been rocked by the COVID-19 pandemic (*Economic Impacts of COVID-19 on Ghana’s Banking Industry - KPMG Ghana*, 2020). Five out of the eight listed banks experienced a fall in their share prices, out of which three experienced a fall of 10% or more for the most part of April, 2020 (*Economic Impacts of COVID-19 on Ghana’s Banking Industry - KPMG Ghana*, 2020).

The hardest hit bank in April was Ecobank Ghana (EGH) falling by 20% from GH¢ 8.10 as at 31^st^ December, 2019 to GH¢ 6.50 as at 21^st^ April, 2020 (*Economic Impacts of COVID-19 on Ghana’s Banking Industry - KPMG Ghana*, 2020). Ghana further closed its air, land and sea borders to human traffic on the midnight of Sunday 22^nd^ March, 2020 resulting in flight cancellations and disruptions (*Ghana Closes Its Borders for 2 Weeks to Fight Coronavirus - Graphic Online*, 2020). Kotoka International Airport in Ghana’s capital city Accra, reopened for regular international passenger travel on Tuesday 1^st^ September, 2020 (*Ghana COVID-19 Information | U.S. Embassy in Ghana*, 2020).

Meanwhile, Ghana’s land and sea borders remain closed until further notice (*Ghana COVID-19 Information | U.S. Embassy in Ghana*, 2020). Ghana’s strategy for managing the COVID-19 pandemic has yielded positive results (*Ghana’s COVID-19 Management Yields Positive Results ∼ Official | CGTN Africa*, 2020). These positive outcomes in the disease management were due to the strategy of early detection, quarantine and treatment the Government of Ghana had adopted from the beginning of the pandemic (*Ghana’s COVID-19 Management Yields Positive Results ∼ Official | CGTN Africa*, 2020).

From the early days of aggressive contact tracing, to quarantine, to mandatory wearing of face mask, washing of hands, use of hand sanitizers and social distancing, a lot more strategies have been an effective strategy for Ghana in battling COVID-19 (*Coronavirus: Is Ghana Winning the Fight against the Pandemic?*, 2020).

### 1.2 Problem Statement

Specimen collection for SARS-CoV 2 testing as it stands currently are through respiratory or mostly pharyngeal specimens [nasopharyngeal / oropharyngeal swab, nasopharyngeal wash, etc] (*Interim Guidelines for Clinical Specimens for COVID-19 | CDC*, 2020). The sampling procedure for these specimens especially nasopharyngeal and oropharyngeal swabs are invasive. It exposes Health workers performing the sampling to a higher risk of contracting SARS-CoV 2 even when in PPEs through aerosolization and is also very uncomfortable or unpleasant to the patients being sampled (*Laboratory Testing for Coronavirus Disease (COVID-19) in Suspected Human Cases*, 2020).

Three thousand (3000) saliva droplet could be generated by one cough which is equal to the amount produced during a five-minutes talk (Xu et al., 2020). Around forty-thousand (40,000) saliva droplets reaching several meters in the air can also be generated by one sneeze (Xu et al., 2020). At the same time, the collection of nasopharyngeal and oropharyngeal specimen types causes minor injuries such as bleeding and ulceration of mucosal layer, especially in patients with predisposing factors (Rao et al., 2020). The sampling technique makes them cough or sneeze profusely during or after sampling (*Alternative Sample Types Could Boost COVID-19 Testing*, 2020). Nasopharyngeal or oropharyngeal swabs that may even be self-collected by suspected COVID-19 patients themselves in other jurisdictions without the necessary assistance from a trained Health worker may not be ideal for SARS-CoV 2 testing (Abdollahi et al., 2020). However, this assistance provided by Health workers conducting the sampling exposes them to contracting SARS-CoV 2 which could eventually result in possible death especially if the infected Health worker has a comorbidity such as hypertension or diabetes (*Association of Medical Laboratory Scientists Loses Member to Covid-19 - MyJoyOnline.Com*, 2020).

This has eventually made the search for an alternative sample with a different sampling procedure such as self-sampling by suspected patients themselves very necessary and urgent in our collective effort globally to defeat the COVID-19 pandemic as well as reduce the number of Health workers who are getting infected in the line of duty (*Alternative Sample Types Could Boost COVID-19 Testing*, 2020).

### 1.3 Justification

Collection of respiratory specimens for SARS-CoV 2 testing is an invasive procedure which causes severe discomfort, mild to intense coughing afterwards by patients (Zhu et al., 2020).

More worrying in particular is the close contact involved between Health care workers sampling the patients and the patients being sampled (*Investigating and Responding to COVID-19 Cases in Non-Healthcare Work Settings | CDC*, 2020). This sampling procedure therefore requires the use of Personal Protective Equipment (PPE) by Health care workers doing these invasive sampling of patients (Park, 2020).

This puts a strain on the scarce PPEs available at a time where there is global shortage of PPEs and even resources to purchase these PPEs in Countries such as Ghana (Park, 2020).

Aside this challenge, the possibility of Scientists and other Health care workers contracting COVID-19 nosocomially because of the close contact involved in these respiratory sampling techniques also exists (Zhu et al., 2020). It has been reported that, in line of duty, some Health workers in Ghana have unfortunately contracted the virus with some even losing their lives but the report could not state scientifically or emphatically that they contracted the virus in the line of duty or from community spread (*Association of Medical Laboratory Scientists Loses Member to Covid-19 - MyJoyOnline.Com*, 2020).

Some studies have made a case for the possible use of saliva or faeces as an alternative sample of choice for COVID-19 testing by RT-PCR because it is non-invasive and easier for patients to produce by themselves (Zhu et al., 2020). This is because the salivary gland and tongue are possibly major sites for replication and viral shedding of SARS-CoV 2 because these tissues express the ACE2 receptor needed for SARS-CoV 2 entry into cells (Rao et al., 2020) as well as the high expression of ACE2 in the mucosal cells of the intestines makes it a potential site for SARS-CoV 2 entry and replication (A. Kumar et al., 2020). It requires no special chilled viral transport media for transportation of samples and there is less specimen degradation resulting from delay in processing (Rao et al., 2020).

Collection of saliva or faecal specimen is done without the help of a Health care worker requiring the use of PPE with its concomitant risk of exposure (*COVID-19 - Control and Prevention | Healthcare workers and Employers | Occupational Safety and Health Administration*, 2020). A similar study in Ghana using Ghanaian COVID-19 patients is therefore urgent and necessary in our collective national effort and global preparedness to defeat COVID-19.

The Ghana Association of Medical Laboratory Scientists (GAMLS) communication outfit have said that over fourteen (14) of its members some of whom include those who were involved in sample taking per data available to them have contracted SARS-CoV 2 across the country (*Coronavirus: 14 Lab Scientists Infected – GAMLS*, 2020). Per a joint press release dated 9^th^ July, 2020 by all the Health Sector Unions and Professional Associations in Ghana, 779 Health workers had contracted SARS-CoV 2 as at 30^th^ June, 2020 with 9 Health workers unfortunately losing their lives (*Health workers Test Positive for COVID-19 in Ghana - Graphic Online*, 2020).

This figure included 190 doctors, 410 nurses & midwives, 23 pharmacists and 156 Allied & other Health workers (*Health workers Test Positive for COVID-19 in Ghana - Graphic Online*, 2020). Among the reasons they stated for this high number of infections among Health workers is the inadequate and erratic supply of PPEs to Health workers both in quality and quantity (*Health workers Test Positive for COVID-19 in Ghana - Graphic Online*, 2020).

Globally, according to the WHO estimate, about 10% of the global SARS-CoV 2 infected people are Health workers with more than 5% of the infected population in 14 Sub-saharan African countries being Health workers (*Over 10 000 Health workers in Africa Infected with COVID-19 | WHO | Regional Office for Africa*, 2020).

Finding an alternative sample and sampling technique will therefore have a positive impact on the Ghanaian, African and Global number of Health workers getting infected with SARS-CoV 2 by reducing these huge numbers of frontline Health workers who are contracting the virus (*Over 10 000 Health workers in Africa Infected with COVID-19 | WHO | Regional Office for Africa*, 2020).

### 1.4 Research Questions

2. Is SARS-CoV 2 shed in the faeces and saliva of confirmed Ghanaian COVID-19 patients?
3. What is the sensitivity and specificity of using faeces in testing for the presence of SARS-CoV 2 as compared to pharyngeal samples (nasopharyngeal/oropharyngeal swabs)?
4. What is the sensitivity and specificity of using saliva in testing for the presence of SARS-CoV 2 as compared to pharyngeal samples (nasopharyngeal/oropharyngeal swabs)?

### 1.5 Study Objectives

#### 1.5.1 General Objective

To verify if saliva or faeces could be a possible good surrogate sample for SARS-CoV 2 RT-PCR testing compared to the current accepted pharyngeal samples (nasopharyngeal/oropharyngeal swabs) in suspected Ghanaian COVID-19 patients attending KATH.

#### 1.5.2 Specific Objectives

1. To ascertain if there is SARS-CoV 2 viral shedding in the saliva and faeces of Ghanaian COVID-19 patients.
2. To assess the sensitivity and specificity of using saliva in determining the presence of SARS-CoV 2 in confirmed COVID-19 patients.
3. To assess the sensitivity and specificity of using faeces in determining the presence of SARS-CoV 2 in confirmed COVID-19 patients.

### 1.6 Study Significance

Current sampling for SARS-CoV 2 testing via RT-PCR is invasive, causes suspected patients being sampled some level of pain, sneezing and coughing with its resultant aerosolization of the immediate sampling environment. This exposes Health workers conducting the sampling to contracting SARS-CoV 2 nosocomially and lastly, is very uncomfortable to the suspected patients being sampled. This study seeks to verify if Ghanaian COVID-19 patients equally sheds the SARS-CoV 2 virus in their saliva and faeces just like some studies in other parts of the world has suggested and whether any of these two samples can possibly replace the current invasive respiratory samples being used for SARS-CoV 2 testing in Ghana.

The findings of this study will benefit suspected COVID-19 patients in terms of the severe discomfort, pain and possible trauma or abrasion they go through with the current invasive respiratory sampling technique. The number of Scientists and other Health care workers getting involved in sample taking for COVID-19 testing with its resultant risk of nosocomially contracting SARS-CoV 2 will be reduced considerably if not eliminated completely. This will preserve the needed staff strength of our hardworking heroes (Health workers) who are at the forefront of this battle against COVID-19.

Personal Protective Equipment (coveralls, N95 nose masks, face shield, googles, gloves), viral transport medium, swab sticks and a host of other consumables needed for respiratory sampling of suspected COVID-19 patients will no more be needed or procured by Government or the Ministry of Health for this purpose. All that will be procured are sterile stool containers which are very cheap and can be used for collecting both the saliva and faecal samples.

This will greatly help the Government of Ghana, Ministry of Health and its Agencies to free some significant amount of resources and possibly re-direct this into providing more testing centers across the country. It will further augment the existing few ones and boost our national preparation towards a possible community surveillance study to know the true prevalence of COVID-19 in Ghana or in preparation towards a possible spike in new SARS-CoV 2 infections in Ghana as is happening currently in other parts of the world.

### 1.7 Framework For SARS-CoV 2 Testing

#### 1.7.1 The Pathway For SARS-CoV 2 Testing Using Nasopharyngeal or Oropharyngeal Swabs/Samples (Gold Standard)

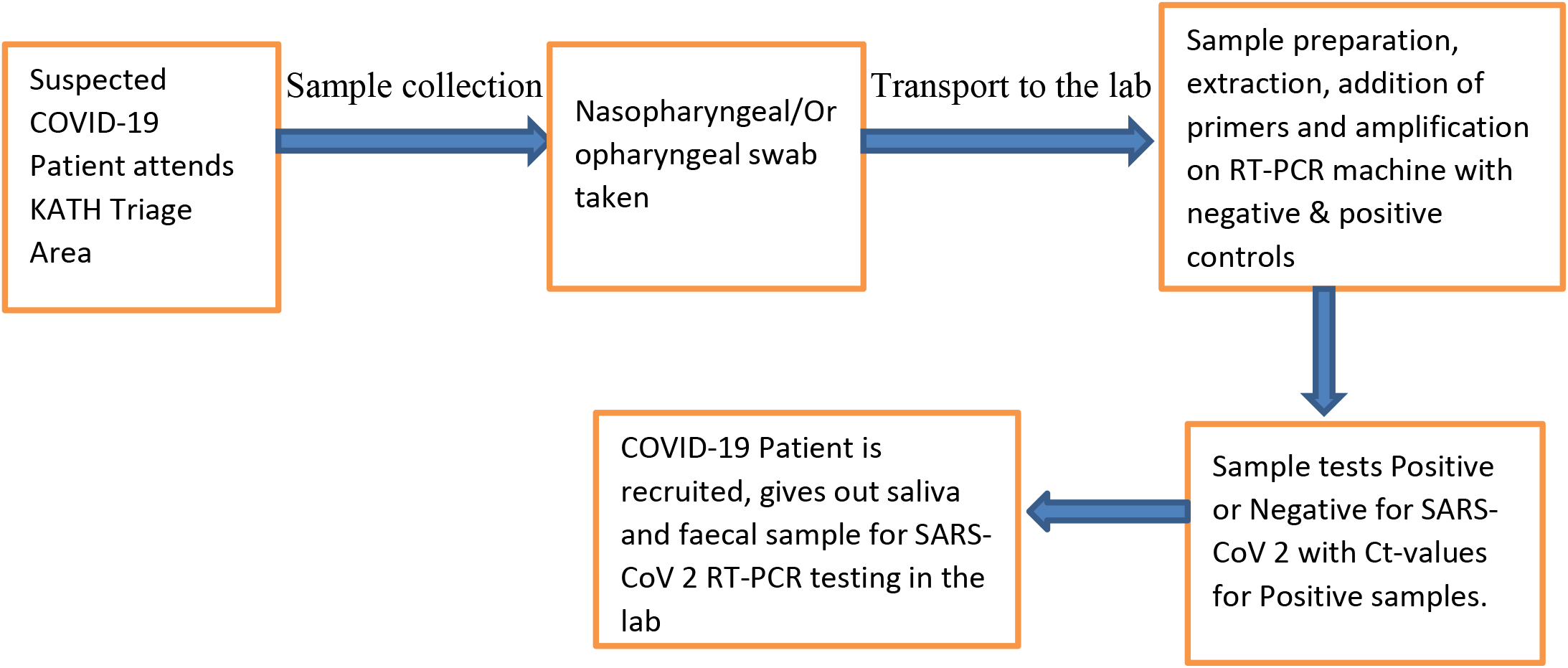

#### 1.7.2 The Pathway For SARS-CoV 2 Testing Using Faecal/Stool Samples

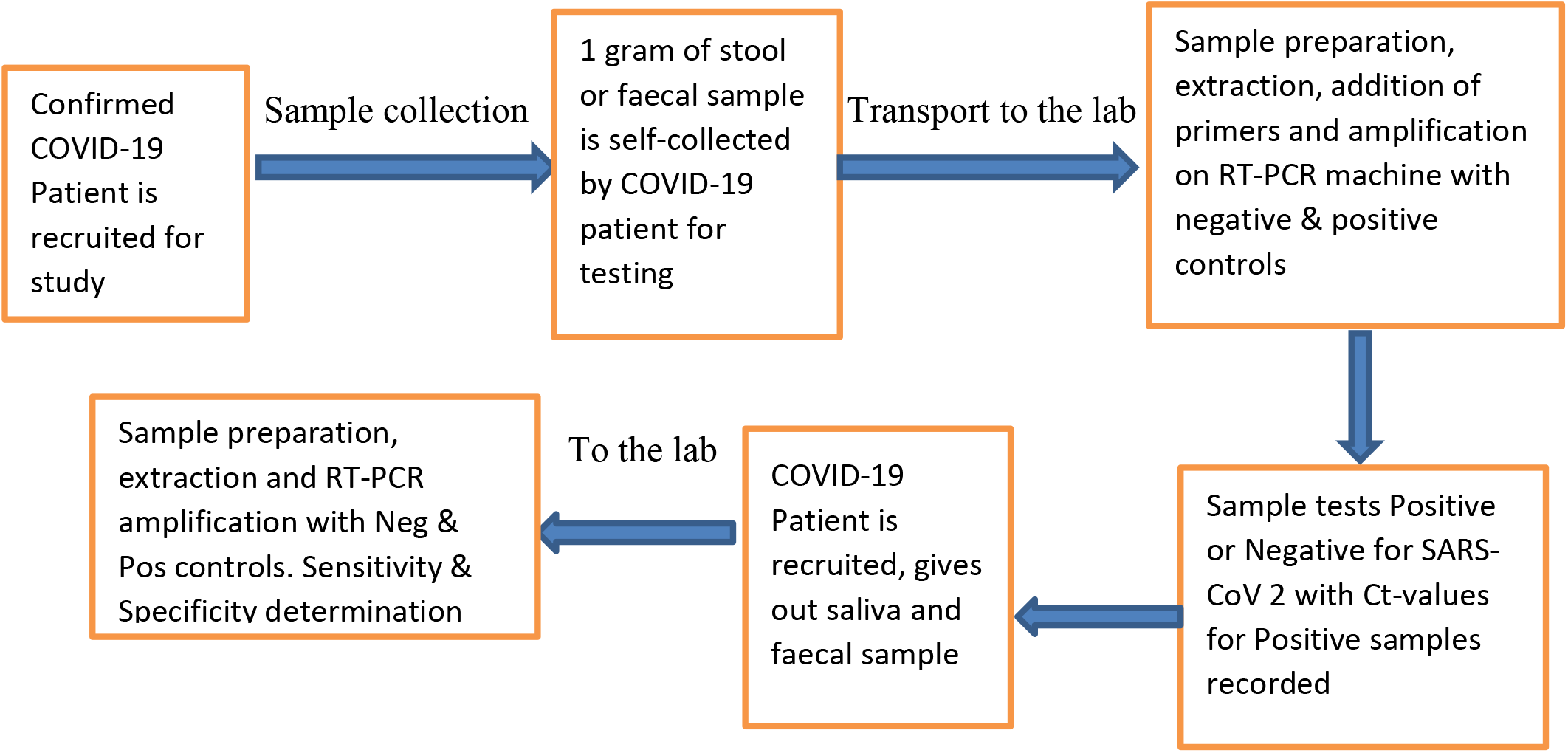

#### 1.7.3 The Pathway For SARS-CoV 2 Testing Using Saliva Samples

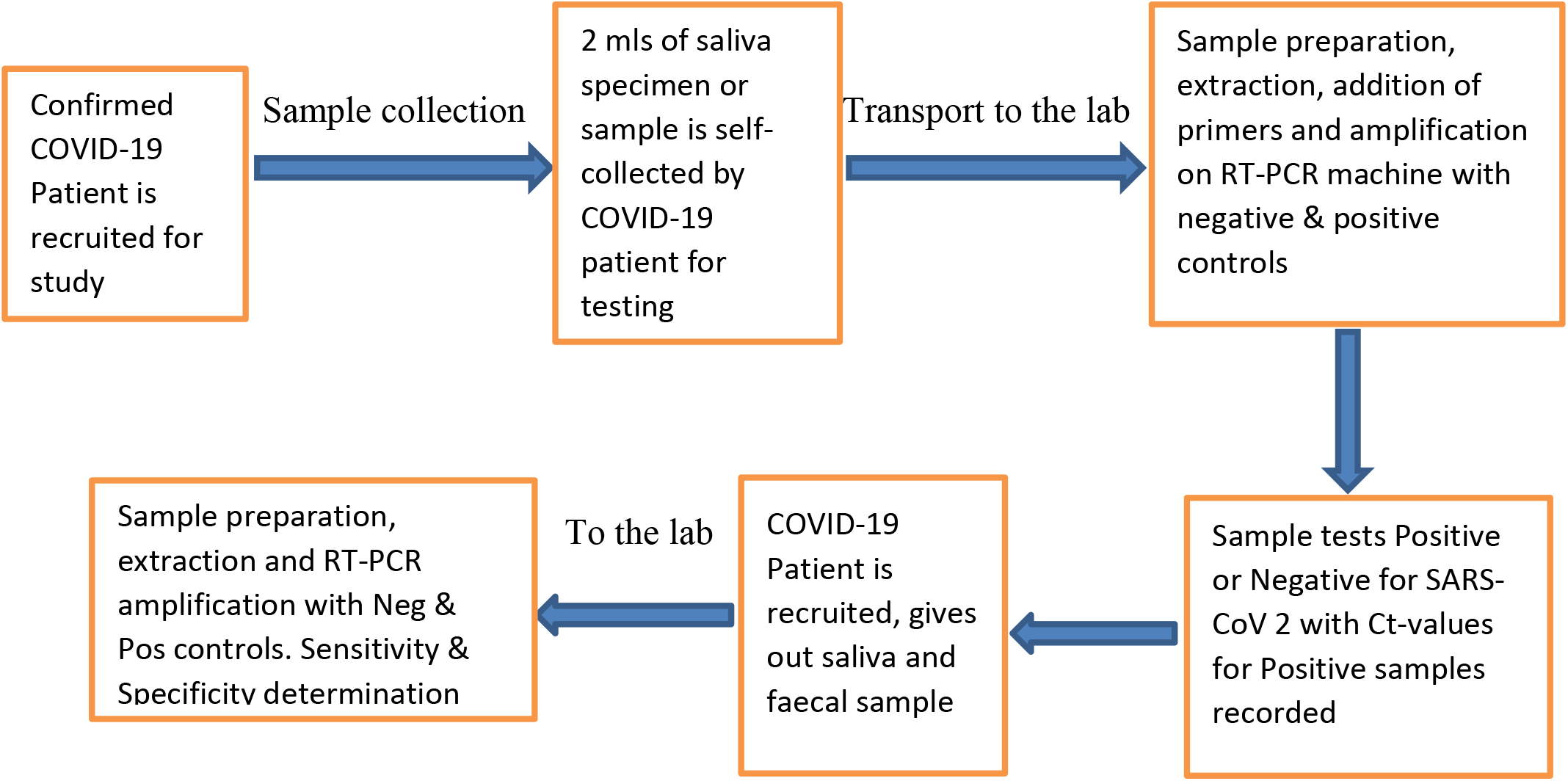

## METHODOLOGY

### 3.1 Background of the Study Area

Komfo Anokye Teaching Hospital is the second largest teaching hospital in Ghana located within Kumasi, the Regional Capital of the Ashanti Region with a population of over four million inhabitants (2010 Ghana Population Census). It is a 1,200-bed capacity hospital and serves as a referral hospital for about twelve out of the sixteen administrative regions in Ghana including Bono, Bono East, Ahafo, Western North, Savannah, Northern, North East, Upper East, Upper West and some parts of the Central and Eastern Regions of Ghana. It also receives patients from neighboring countries such as Ivory Coast and Burkina Faso.

In the year 1940, the British Colonial Government built an African and European Hospital over a small hill over-looking the Bantama Township with the African side treating African patients and the European side treating Europeans who were living in Kumasi at the time. The need to construct a bigger hospital to cater for the growing population of Kumasi came in 1952. This led to the transfer of the European Hospital to the Kwadaso Military Quarters to pave way for the new hospital complex. So, by 1954, British contractors by name Gee, Walker & Slater have completed this new hospital complex which named Kumasi Central Hospital with commencement of operation in 1955.

The name was later changed to Komfo Anokye Hospital in honour and memory of Okomfo Anokye, a powerful and legendary fetish priest of Asanteman. However, local inhabitants have popularly referred to the hospital as “Gee”, naming it after one of the British contractors that built the Hospital in 1954 and this local name have remained till today. It became a Teaching Hospital in 1975 following the establishment of the School of Medical Sciences in Kwame Nkrumah University of Science and Technology (KNUST).

The Hospital currently have a workforce of over four thousand (4000) workers with varied professional backgrounds offering both clinical and non-clinical services with about 13 clinical directorates and 2 non-clinical directorates.

The clinical directorates include Emergency Medicine, Surgery, Trauma and Orthopaedics, Medicine, Obstetrics and Gynaecology, Child Health, Family Medicine, Laboratory Services, Oncology, EENT (Eye, Ear, Nose & Throat), Radiology, Oral Health and the Anesthesia & Intensive Care Directorate. The two non-clinical directorates are Domestic and Technical Services Directorates. There are also a number of Units including Public Health, Transfusion Medicine, Information Technology (ICT), Environmental, Human Resource, General Administration, Biostatistics and Security Units.

The Vision of the Hospital is to become a center of excellence in the provision of specialist healthcare services and its mission is to provide quality healthcare services to meet the needs and expectations of all its patients. This they hope to achieve through a well-motivated and committed staff applying best practices and innovativeness. The Hospital is currently one of the centers for the treatment of COVID-19 patients in Ghana with the current on-going global pandemic. It has an Infectious Disease Isolation Area (IDHA) and a Highly Infectious Isolation Unit (HIIU) which was all created following the declaration of the global pandemic to receive patients who may need treatment from COVID-19.

### 3.2 Study Design and Type

This study is an experimental case study with focus on COVID-19 patients faecal and saliva viral shedding aimed at generating some preliminary data to be able to make some inferences about the possible use of saliva or faeces for COVID-19 testing instead of the current invasive pharyngeal sampling or samples being used.

Patients who were confirmed as having tested positive for SARS-CoV 2 using their nasopharyngeal or oropharyngeal swabs as well as those who were suspected COVID-19 patients but tested negative for SARS-CoV 2 using same sample were recruited as study participants.

Confirmed COVID-19 patients as well as suspected patients who tested negative for SARS-CoV 2 and eventually became study participants after consenting to be part of this study were sampled once for their saliva and faecal sample which was analyzed via RT-PCR using same reagent as was used for their pharyngeal sample testing. There was no follow up sampling or repeated sampling of the participants at different times and dates.

### 3.3 Study Population

The study population were suspected COVID-19 patients who were on admission at the Infectious Disease Holding Area (IDHA) of the Komfo Anokye Teaching Hospital (KATH) after meeting the case definition protocol and have tested positive for SARS-CoV 2 by RT-PCR using nasopharyngeal/oropharyngeal swabs. Confirmed COVID-19 Patients who were also on admission at the Highly Infectious Isolation Unit (HIIU) located within the premises of the Family Medicine Directorate (FMD) were also sampled.

Patients who reported with mild symptoms of COVID-19, tested positive for SARS-CoV 2 and decided or were given the option by Healthcare professionals to undergo home management and isolation instead of being on admission were also contacted and those who agreed to provide a sample were recruited with their consent. Every suspected COVID-19 patient that visited KATH and tested positive for SARS-CoV 2 via RT-PCR with pharyngeal sample had an equal chance of getting recruited to be part of this study. Some suspected COVID-19 patients who tested negative for SARS-CoV 2 with their respiratory samples were also recruited to be part of this study for the purpose of meeting the specific objective 2 and 3 that has to do with the determination of the sensitivity and specificity of using saliva or faeces as compared to the pharyngeal samples (gold standard).

### 3.4 Sampling Technique and Sample Size

The Convenience Sampling Method which is a non-probability sampling method was employed in this study to recruit participants. This is because the study sought to find some preliminary data as a basis for a larger study to be undertaken in future. Saliva and Faecal samples were collected from all confirmed COVID-19 patients whose pharyngeal swab sample already taken tested positive for SARS-CoV 2 by qualitative RT-PCR and consented to be part of this case study through a signed consent form that was read and administered to them. Some suspected COVID-19 patients who tested negative for SARS-CoV 2 and consented to be part of this study were also recruited.

After consenting and successful recruitment as a study participant, two sets of sterile stool containers were given to each recruited patient or participant and they were asked to supply about 1gram of stool/faecal sample and 1-2 mls of early morning saliva by themselves using standard stool and saliva collection technique used by every other patient. Samples collected were quickly triple packaged and transported to the KATH Infectious Disease and Research Laboratory (IDRL) and processed in order to preserve their viability and integrity.

Patients who tested positive for SARS-CoV 2 and were at their various homes isolating and treating themselves were equally contacted and recruited. Once they consented and got themselves recruited, two sterile stool containers were sent to them at their places of abode for them to provide about 1 gram of stool and 1–2 mls of early morning saliva the following morning. Once they were done with the sampling, they placed a call and the samples were quickly picked up and immediately sent to the IDRL for processing.

The suspected COVID-19 patients who tested negative for SARS-CoV 2 and were mostly at their various homes were also contacted and those who consented were recruited and sampled just like the other recruited patients.

Asymptomatic patients who reported to KATH voluntarily to test for SARS-CoV 2 either as a requirement for international travel or medical examination purposes and tested positive were all recruited and those who consented to be part of this study were sampled just like the others as well.

The sample size or study participants that were successfully recruited to be part of this study totaled fifty (50) confirmed COVID-19 patients and twenty (20) suspected COVID-19 patients whose pharyngeal samples tested negative for SARS-CoV 2. This brought the total number of patients recruited to seventy (70) participants. This culminated into three (3) RT-PCR tests which was conducted on each patient using the three (3) different samples (pharyngeal, saliva and faeces) with a total of two-hundred and ten (210) RT-PCR tests done for this study. The patients or participants recruitment exercise took place between October – December, 2020 at KATH.

### 3.5 Study Variables

The study variables included the Dependent and Independent variables. The Dependent variable in this study was viral shedding in the alternate specimens (saliva and faeces) taken from the recruited patients. Independent variables in this study included factors that had the potential to influence the Dependent variable and may affect viral shedding of SARS-COV 2 in saliva or faecal samples of Ghanaian COVID-19 patients or study participants.

They include the sampling time especially the saliva sample, number of days after symptoms onset and medication that the patients may have taken prior to sampling at KATH which may all affect viral shedding in the sample of interest.

Another point is that, ACE2 messenger RNA is highly expressed and stabilized by the presence of a neutral amino acid transporter in the gastrointestinal system. So, anything that can affect the presence or function of the neutral amino acid transporter will also affect viral shedding in faecal samples.

Another independent variable is the possible activation of SARS-CoV 2 virus by gene expression of furin in salivary glands of patients. So, without furin fully expressing itself in the salivary gland due to any condition affecting it, SARS-CoV 2 viral shedding in saliva could also be affected.

### 3.6 Data Collection Tool and Technique

Demographic data on each recruited patient (age, sex, type of symptoms, district/municipal or metropolis and region) were extracted from the Case Investigation Form for suspected COVID-19 patients that accompanied the pharyngeal samples brought for testing at the Infectious Disease and Research Laboratory (IDRL).

Laboratory analysis of all the pharyngeal, saliva and faecal samples taken from each recruited patient also provided critical data on the presence or absence of SARS-CoV 2 in each of the samples as well as the determination of their cycle time or threshold (Ct) values as well as their sensitivity and specificity using the true positive and negative rates. The raw data was inputted and stored in an excel sheet. The raw data was generated through the qualitative Real Time-Polymerase Chain Reaction (RT-PCR) for the detection of SARS-CoV 2 from pharyngeal, saliva and faecal specimens. The following consumables and equipment were used throughout the process of data generation from sample collection to detection or otherwise of SARS-CoV 2 from the samples via RT-PCR;

i. Pipette Tips (1000uL, 200uL & 10uL) manufactured by Henso Medical Company Ltd, Hangzhou, China.
ii. Pharyngeal swab sticks, tongue depressors, viral transport medium, N95 nose mask, eye goggles and coveralls manufactured by Jiangsu Rongye Technology Co., Ltd. Jiangsu, China.
iii. Saliva & Stool containers by Jiangsu Huida Medical Instruments Company Ltd, Jiangsu-China.
iv. Triple package transport container manufactured by ULAB Scientific, Nanjing, China.
v. Microcentrifuge Tubes (2.0ml & 1.5ml) manufactured by Changde Bkmam Biotechnology Company Ltd, China.
vi. Rota Mixer manufactured by Hook & Tucker Instruments Ltd, Croydon-United Kingdom.
vii. Thermo Heating block (Dry Bath) assembled in China by Nanjing Binzhenghong Instrument Co., Ltd.
viii. Microcentrifuge (Micro 200) Zentrifugen manufactured by Sigma Laboratory Centrifuges, Germany.
ix. Telstar BV – 100 Biosafety Cabinet BSL II manufactured by Telstar Laboratory Equipment BV based in Woerden, Netherlands.
x. Extraction Kit (Nucleic Acid Extraction and Purification Reagents), Series C (Spin column method) manufactured by Guangzhou LPB Medicine Science & Technology Company Ltd, China.
xi. Dragon Lab Micropipettes (10uL, 1000uL, 100uL, 200uL & 50uL) manufactured by Sri Sai Plastics Enterprises in Lucknow, Uttar Pradesh-India.
xii. Nitrile powdered Examination gloves and disposable vinyl examination gloves manufactured by Chagnzhou TTA Medical Instrument Co.,Ltd, Wujin District, Changzhou, Jiangsu Jiangsu-China.
xiii. Real Time-Polymerase Chain Reaction Machine manufactured by Agilent Technologies Stratagene Mx 3005p based in Santa Clara, California-USA.
xiv. Detection Kit for 2019-Novel Coronavirus (PCR-Fluorescence probing) by Da An Gene Company Ltd of Sun Yat-Sen University in Guangzhou, GuangDong Province, China P.R.

#### 3.6.1 Procedure for The Extraction and RT-PCR Testing For SARS-CoV 2 Using Pharyngeal Swabs, Saliva and Faecal Samples From Study Participants

With nasopharyngeal or oropharyngeal swabs, 0.9% saline or normal saline was added to wash the swab to obtain a viral solution or wash which was used for the extraction of viral RNA. For saliva samples, 0.9% sterile saline solution was added to wash the saliva and get a viral solution. For the Faecal/Stool samples, 0.9% sterile saline was first added, mixed well and centrifuged at 2,500rpm for 5 minutes.

The supernatant which was in the form of a wash and contains the virus was then aliquoted and used for the viral extraction process. All three samples of each participant were subjected to the same RT-PCR testing process after this initial preparation to obtain the viral solutions.

The reagent kit used for the viral RNA extraction, enrichment and purification of all three samples (pharyngeal, saliva and faeces) prior to the RT-PCR testing was the *LBP Viral RNA Mini Kit*. Before the SARS-CoV 2 Viral RNA extraction process was started, the following essential activities were first carried out;

i. Dry-bath was switched on and temperature set to 72^0^ celcius.
ii. Working bench surfaces were decontaminated together with all applicable equipment with 70% alcohol.
iii. All reagents removed from storage for use were allowed to equilibrate to room temperature.
iv. 100ul of elution buffer was aliquoted per sample and incubated at 72^0^ celcius.
v. Isopropanol or ethanol was added into a Lysate and content added to wash solution 1 & 2 bottles and labelled.
vi. The Elution buffer was pre-heated at a temperature of 72^0^ celcius.
vii. All participant samples were given unique identity numbers to avoid sample mix up.

After these essential activities, the SARS-CoV 2 viral RNA extraction process was then performed through the following processes;

i. 400uL of Lysate (with isopropanol added) was taken with a pipette into a 1.5mL centrifuge tube.
ii. 200uL of the sample in the form of a viral solution was then added to the Lysate in the centrifuge tube.
iii. 20uL of Proteinase K was then added to the content in the centrifuge tube and vortexed for 10 seconds to form a uniform mixture. The mixture was then incubated at 72^0^ celcius for 10 minutes using the dry temperature heater to lyse the virus.
iv. The solution was then transferred into spin columns and centrifuged at 1,200rpm for 60 seconds. This allows the viral nucleic acid to bind to the columns.
v. The filtrate was then discarded, spin column was put back into centrifuge tube, 500uL of wash solution 1 was added and centrifuged at 1,200rpm for 60 seconds.
vi. The filtrate was discarded once again, spin column was put back into centrifuge tube and another 500uL of wash solution 1 added again as well as centrifuged for 60 seconds at 1,200rpm.
vii. The filtrate was discarded once again, spin column put back into centrifuge tube, 500uL of wash solution 2 was then added and centrifuged at 1,200rpm for 60 seconds.
viii. The filtrate was discarded, spin column was put back into its original collection tube and centrifuged at 1,200rpm for 3 minutes. This is dry spinning.
ix. The spin column was then transferred to a new 1.5mL centrifuge tube, incubated at 72° celcius for 2 minutes. 50ul of elution buffer was then added, mixture was allowed to stand at room temperature for 60 seconds and then centrifuged at 12,000rpm for 60 seconds. The elusion buffer dislodges the viral RNA from the columns.
x. The extracted SARS-CoV 2 nucleic acid needed for the RT-PCR were present in the elution buffer at this stage after centrifugation. It was then used for the RT-PCR amplification. Extracted nucleic acid not immediately used for RT-PCR analysis was stored at −80° celcius.

At the RT-PCR room, the airconditioner was set to 16^0^celcius before the PCR Machine (Strategene Mx3005P) was switched on. The probes that were used included the FAM (Test), HEX (Test) and CY5 (Internal Control). The following procedures led to the final stage of testing the extracted nucleic acid via RT-PCR;

i. The Strategene Mx3005P RT-PCR Machine was switched on.
ii. The MxPro Mx3005P quantitative PCR software on the computer connected to the PCR machine is launched.
iii. Number of wells were highlighted according to number of samples to be run.
iv. “Unknown” was selected from the well type drop down button.
v. The check boxes were clicked for FAM, HEX and CY5 probe detection channels and “show well names” button was also clicked on the computer.
vi. The first highlighted well was right clicked and the button “well information” was selected.
vii. The well or sample identity for each participants sample were then inputted and “Thermal profile” was clicked.
viii. Previous segments were deleted and “segment” was clicked 3 times with the last segment highlighted and “plateau with ramp” selected or clicked.
ix. The various temperature and time for each segment were inputted as follows to form the Thermal Cycle Profile below;
x. The assay was then saved and the start button was clicked on the computer for the RT-PCR analysis to begin. Checkbox for “Switch off Lamp after test” was clicked on the “Run status panel”.

**Table 1:**
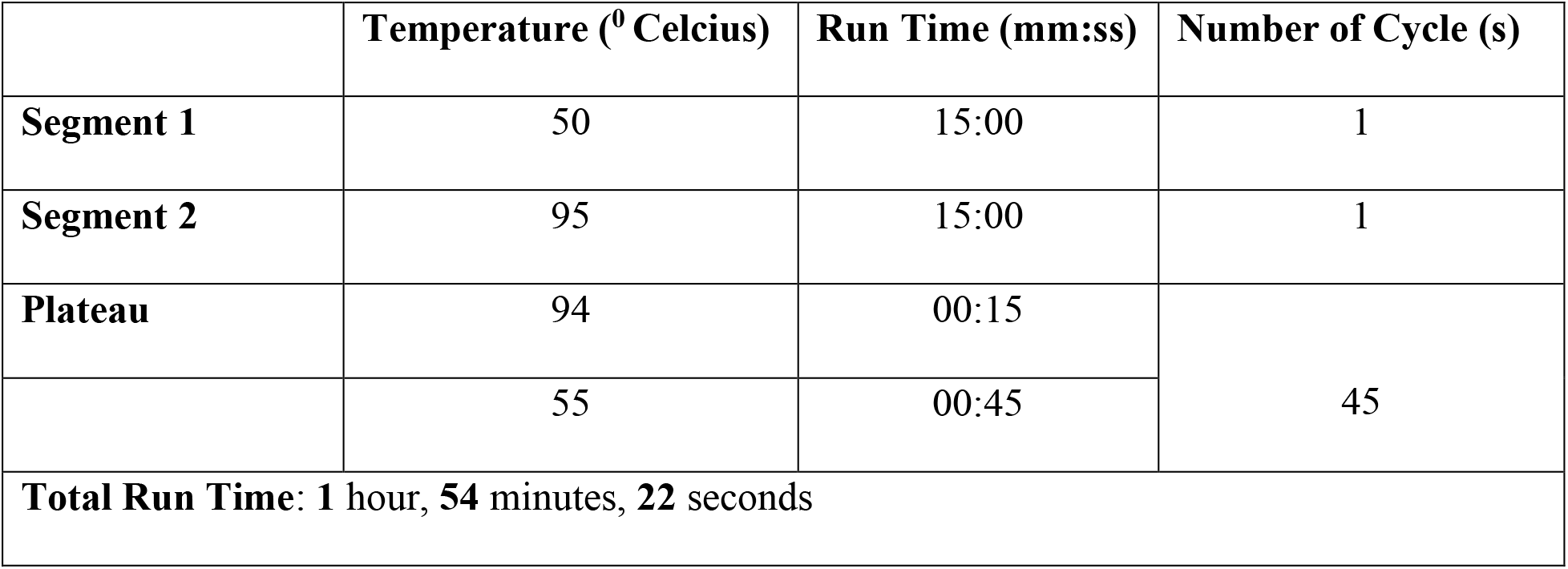
RT-PCR Analysis Parameters.

In the analysis mode, the amplification graph was clicked and Cycle threshold (Ct) for each probe was set. To view results for each participant sample, “plate value” was clicked. The results were interpreted as follows;

1. Participants whose sample had number of Ct value greater than 40 in the FAM and HEX channels with an amplification curve in the CY5 channel were declared ***NEGATIVE*** for SARS-CoV 2.
2. Participants whose sample had a Ct value of 40 and below for the FAM and HEX channels were declared ***POSITIVE*** for SARS-CoV 2.
3. Test was repeated if only one of the FAM or HEX Ct values is 40 or less and the other is greater than 40. If the retest results come out same as the previous test, that sample was taken as ***POSITIVE*** for SARS-CoV 2.

### 3.7 Data Analysis

Data obtained from this study were analyzed using R analytics software, a programming language for statistical computing and graphics that is normally used to clean, analyze and graph data. It is widely used by researchers from diverse disciplines to estimate and display results data analysis. Demographic data, RT-PCR results from the pharyngeal, saliva and faecal samples were all analyzed by subjecting them to both descriptive as well as inferential statistical analysis with graphs that will lead to some inferences being suggested. The sensitivity and specificity of saliva and faeces being used for SARS-CoV 2 RT-PCR testing as against the gold standard of pharyngeal specimens were also determined.

### 3.8 Limitation of the Study

This study was influenced by the sampling time of the saliva samples especially. Research have shown that early morning saliva before oral health is very good a sample for SARS-CoV 2 testing in asymptomatic and symptomatic patients as well because of its very high viral presence at that time. Viral particles however reduce after oral health, breakfast and other eating or drinking activities.

So, samples taken in the course of the day could have reduced SARS-CoV 2 viral particles and as such have an effect on the testing. The sample quality of the saliva samples is also another limiting factor especially taking the sample right after eating or drinking water or any fluid affects the sample quality at that very moment.

Delays in sampling some of the asymptomatic patients especially the travelers, medical exams and voluntary patients also affected their sample quality. Some symptomatic patients were already on treatment before sampling. Drugs such as Ivermectin reduces viral load of SARS-CoV 2 and as such may have an effect on viral shedding in saliva or stool samples. Lastly, sample size for this study will also not allow for findings to be generalized which is a limitation.

The ACE2 messenger RNA is highly expressed and stabilized by the presence of a neutral amino acid transporter in the gastrointestinal system. So, conditions such as Hartnup disease or disorder that affects this neutral amino acid transporter will equally have an effect on SARS-CoV 2 viral shedding in stool samples of participants. The possible activation of SARS-CoV 2 by gene expression of furin in salivary glands is also another limitation as furin inhibitors such as Decanoyl-RVKR-chloromethylketone (CMK) and Naphthofluorescein affects the gene expression of furin. They show antiviral effects on SARS-CoV-2-infected cells by decreasing virus production and cytopathic effects.

Whiles CMK blocks virus entry with further suppression of the cleavage of spikes and the syncytium, Naphthofluorescein acts primarily by suppressing viral RNA transcription. This may affect viral shedding in saliva samples of study participants should any of them have these furin inhibitors at the time of sampling.

### 3.9 Ethical Consideration

Ethical clearance was sought or obtained from the Komfo Anokye Teaching Hospital (KATH) Institutional Review Board (KATH-IRB) through the Research and Development (R&D) Unit of the hospital. This was an expediated priority ethical review because of the nature of the study and its relationship with COVID-19 patients in the midst of a pandemic. There was no potential risk or discomfort to any of the study participants as the sample of interest was obtained through a non-invasive, self-sampling procedure that the patients easily carried out themselves after educating them on how to take the samples correctly.

There was no discomfort to the study participants or no participant complained of being discomforted as these two samples (saliva and faeces) were easily produced within the day after consent. Confidentiality was guaranteed as all patients or participants recruited remained anonymous.

The consent form was in simple English which was self-read and understood by most literate participants whiles it was interpreted in the local language for illiterate patients by my study collaborators before they signed or thumb printed the form to consent. No patient suffered any consequence whatsoever being it the way they were medically treated or cared for following their decision to participate in this study or not.

There were no financial or material incentives given to any study participant after consent or producing the samples of interest but all participants were made aware of the benefits that society at large will get from this study should they volunteer to participate.

## RESULTS

This section presents findings from data collected to verify if saliva and faecal specimens could possibly be surrogates for the presently used pharyngeal swabs. The results are presented in tabular forms and graphical representation. In all, 50 patients who tested positive for SARS-CoV2 and 20 patients who tested negative were used as sample for the study.

**Table 2.** Characteristics of Study Participants tested via Pharyngeal Swabs.

|  | **Suspected SARS-CoV2** |  |  |
| --- | --- | --- | --- |
|  | **Nasopharyngeal**<br>**Positive**<br>**N(%)** | **Nasopharyngeal**<br>**Negative**<br>**N(%)** | **Total** |
| **Gender** |  |  |  |
| Males | 22(44) | 10(50) | 32(45.7) |
| Females | 28(56) | 10(50) | 38(54.3) |
| **Age** |  |  |  |
| 10 – 19 | 2 (4) | 0 (0) | 2 (2.9) |
| 20 – 29 | 12 (24) | 3 (15) | 15 (21.4) |
| 30 – 39 | 17 (34) | 9 (45) | 26 (37.1) |
| 40 – 49 | 8 (16) | 4 (20) | 12 (17.1) |
| 50 – 59 | 3 (6) | 2 (10) | 5 (7.1) |
| 60 – 69 | 3 (6) | 0 (0) | 3 (4.3) |
| 70 – 79 | 4 (8) | 1 (5) | 5 (7.1) |
| 80 – 89 | 1 (2) | 1 (5) | 2 (2.9) |
| **Place of residence** |  |  |  |
| Urban | 34 (68) | 14 (70) | 48 (68.6) |
| Sub-urban | 16 (32) | 6 (30) | 22 (31.4) |
| **Symptoms** |  |  |  |
| General Malaise | 1 (2) | 0 (0) | 1 (1.4) |
| General Weakness | 11 (22) | 4 (20) | 15 (21.4) |
| Cough | 25 (50) | 18 (90) | 43 (61.4) |
| Anosmia | 13 (26) | 0 (0) | 13 (18.6) |
| Fever | 16 (32) | 6 (30) | 22 (31.4) |
| Chills | 11 (22) | 3 (15) | 14 (20.0) |
| Chest pains | 5 (10) | 4 (20) | 9 (12.9) |
| Shortness of breath | 8 (16) | 0 (0) | 8 (11.4) |
| Stuffy nose | 1 (2) | 0 (0) | 1 (1.4) |
| Headache | 11 (22) | 5 (25) | 16 (22.9) |
| Sore Throat | 5 (10) | 2 (10) | 7 (10.0) |
| Runny nose | 3 (6) | 1 (5) | 4 (5.7) |
| Loss of Appetite | 1 (2) | 1 (5) | 2 (2.9) |
| Nausea | 1 (2) | 1 (5) | 2 (2.9) |
| Common cold | 1 (2) | 0 (0) | 1 (1.4) |
| Diarrhoea | 1 (2) | 0 (0) | 1 (1.4) |
| Bodily pains | 1 (2) | 0 (0) | 1 (1.4) |
| Asymptomatic | 7 (14) | 0 (0) | 7 (10.0) |

Out of the seventy (70) participants who were successfully recruited for this experimental case study, thirty-eight (38) were females representing 54.3% and thirty-two (32) were males representing 45.7%. The female participants had their ages ranging from 20 – 78 years with a mean age of 41.5 years. The male participants had their ages ranging from 18 – 83 years with a mean age of 38.8 years. Out of the fifty (50) COVID-19 or SARS-CoV 2 positive patients that were successfully recruited for this study, twenty-eight (28) were females representing 56% whiles twenty-two (22) were males representing 44%. Of the twenty-eight (28) SARS-CoV 2 positive females, twenty-two (22) representing 79% approximately were below sixty (60) years of age with their ages ranging from 20 – 56 years whiles six (6) representing 21% approximately were above sixty years of age with their ages ranging from 65 – 84 years.

The mean age of all twenty-eight (28) female SARS-CoV 2 positive participants was 42.9 years. Of the twenty-two (22) SARS-CoV 2 positive male participants, twenty (20) of them representing 91% approximately were below sixty (60) years of age with their ages ranging from 18 – 50 years whiles only two male SARS-CoV 2 positive participants representing 9% were above sixty years of age (65 & 78years). The mean age of all twenty-two (22) SARS-CoV 2 positive males was 36.2 years.

The twenty (20) suspected COVID-19 patients who tested negative and were recruited for this study comprised of ten (10) males and ten (10) females. The mean age of the ten males is 44.6 years with their ages ranging from 23 – 83 years whiles the ten females have a mean age of 37.7 years with their ages ranging from 23 – 56 years.

Among the symptoms experienced mostly by participants included cough (43 patients), Fever (22 patients), chills (14 patients), General weakness (15 patients), chest pain (9 patients) and asymptomatic patients were seven (7).

Other symptoms included general malaise, anosmia, shortness of breath, stuffy nose, headache, sore throat, runny nose, loss of appetite, nausea, common cold, diarrhoea and bodily pains. Fifty-two (52) participants out of the seventy (70) recruited representing 74% resided in an Urban center whiles eighteen (18) representing 26% resided in a Sub-urban center. Sixty-eight (68) participants representing 97% were residing in Ashanti region whiles two (2) representing 3% resided outside Ashanti region, specifically Eastern and Greater Accra region.

### 4.2 Viral Shedding of SARS-CoV 2 in the Saliva and Faeces of Ghanaian COVID-19 Patients

Per the results obtained from this study, out of fifty (50) SARS-CoV 2 positive patients who were confirmed as positive using the gold standard pharyngeal samples, forty-three (43) of them representing 86% had their saliva samples testing positive for SARS-CoV 2 using same RT-PCR procedure and test kit. On the other hand, all fifty (50) stool/faecal samples taken from the study participants who were confirmed COVID-19 patients also came out positive for SARS-CoV 2 (100%) via same RT-PCR with same test kit.

**Figure 3:**
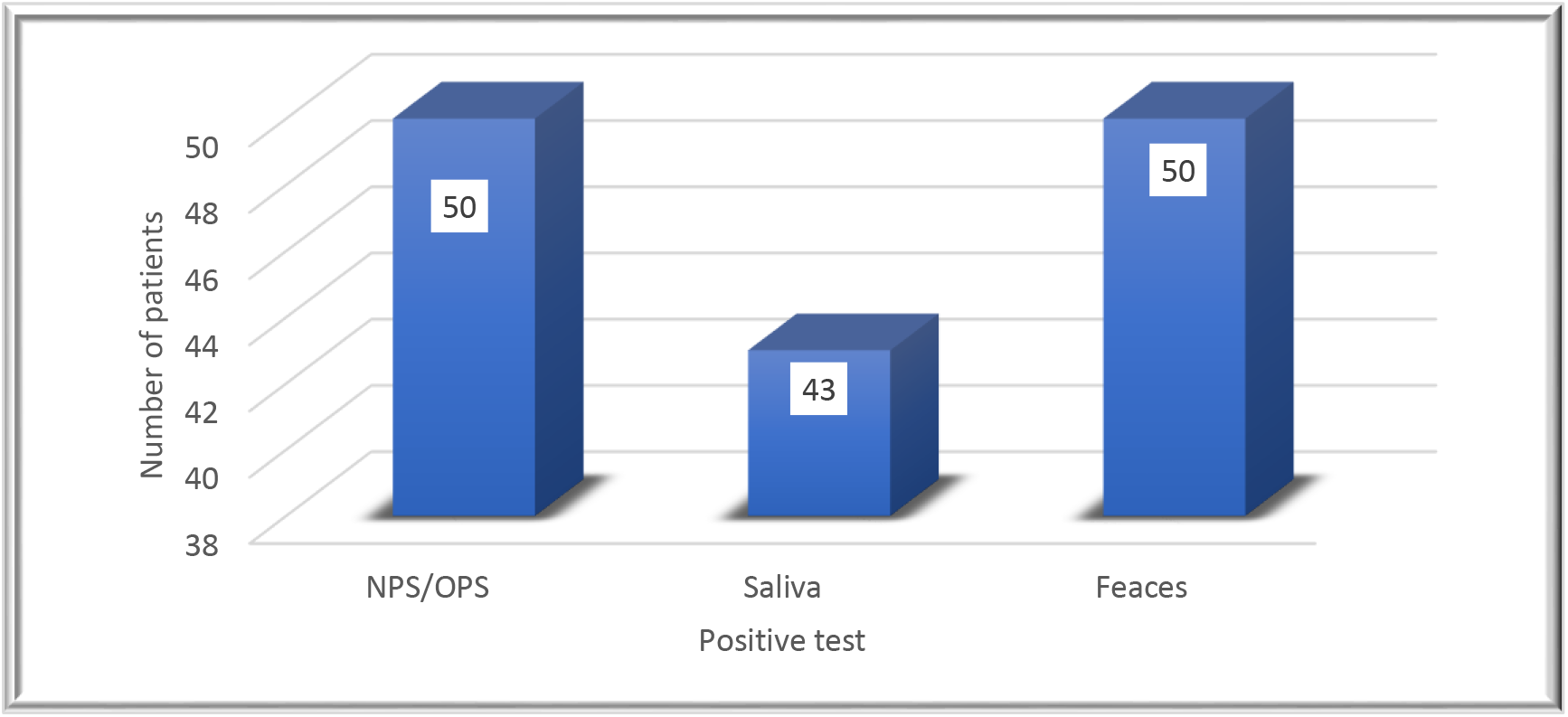
Bar Chart showing the number of positive SARS-CoV 2 participants identified by each sample.

### 4.3 Sensitivity and Specificity of using Saliva to test for SARS-CoV 2 as compared to Pharyngeal samples (Gold standard)

#### Nasopharyngeal/Oropharyngeal Swab and Saliva Sample Results

**Table 3:**
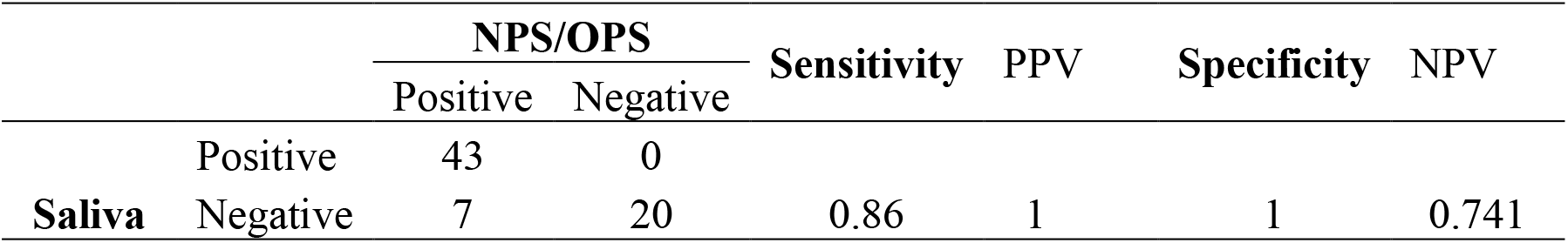
Sensitivity and Specificity of saliva samples as compared to pharyngeal samples.

The sensitivity test results indicated that nasopharyngeal/oropharyngeal swab (gold standard) identified fifty (50) people as having SARS-CoV 2 (positive) via RT-PCR whiles the saliva sample test via same RT-PCR was able to identify 86% of the positive individuals with SARS-CoV 2 as having the COVID-19 Disease. So, the sensitivity of using saliva sample to test for SARS-CoV 2 as against the pharyngeal samples per this study was 86%.

The Positive Predictive Value (PPV) of 100% shows that all the participants who tested positive for SARS-CoV 2 using their saliva sample were among the participants having SARS-CoV 2 when tested using their pharyngeal samples (gold standard).

The specificity results of 100% suggested that all the participants who tested negative for SARS-CoV 2 using their saliva sample test were not having SARS-CoV 2 when tested with their pharyngeal samples (gold standard). However, the Negative Predictive Value (PPV) of 74.1% indicates that 74.1% of participants who tested negative using their saliva samples are truly negative for SARS-CoV 2. So, the False Negatives were seven (7) participants representing 25.9%.

### 4.4 Sensitivity and Specificity of using Faeces to test for SARS-CoV 2 as compared to Pharyngeal samples (Gold standard)

#### Nasopharyngeal/Oropharyngeal swab and Faecal sample results

**Table 4:**
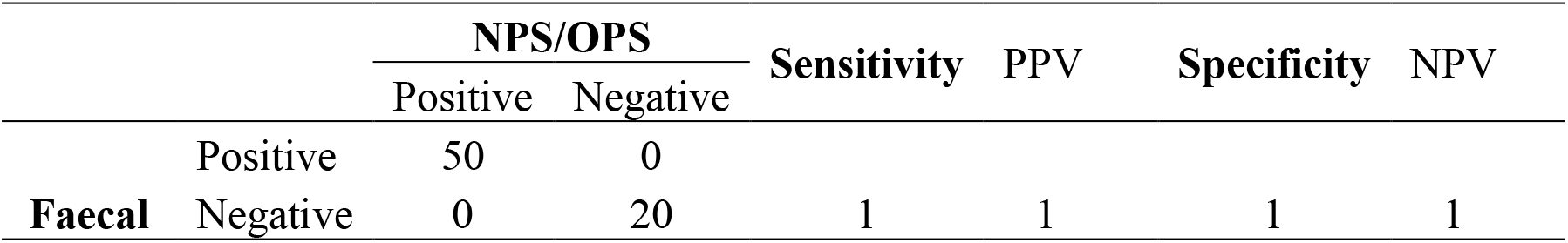
Sensitivity and Specificity of faecal samples as compared to pharyngeal samples.

The sensitivity test results indicated that nasopharyngeal/oropharyngeal swab (gold standard) identified fifty (50) people as having SARS-CoV 2 (positive) via RT-PCR whiles the faecal sample test via same RT-PCR with same test kit was also able to identify 100% of the positive individuals with SARS-CoV 2 as having the COVID-19 Disease.

So, the sensitivity of using faecal sample to test for SARS-CoV 2 as against the pharyngeal samples per this study was 100%. The Positive Predictive Value (PPV) of 100% shows that all the participants who tested positive for SARS-CoV 2 using their faecal sample were actually among the participants having SARS-CoV 2 when tested using their pharyngeal samples (gold standard). The specificity result of 100% suggests that all the participants who tested negative for SARS-CoV 2 using their faecal sample for the RT-PCR test were not having SARS-CoV 2 when tested with their pharyngeal samples (gold standard).

Therefore, the Negative Predictive Value (NPV) of 100% indicates that 100% of all the participants testing negative using their faecal samples are truly negative for SARS-CoV 2. So, there were no False Negatives (0%) so far as using faecal samples to test for SARS-CoV 2 via RT-PCR in this study was concerned.

### 4.5 Sensitivity and Specificity of using Saliva to test for SARS-CoV 2 as compared to Faecal samples

#### Saliva sample and Faecal sample results

**Table 5:** Sensitivity and Specificity of saliva samples as compared to faecal samples.

|  | Saliva |  | Sensitivity | PPV | Specificity | NPV |
| --- | --- | --- | --- | --- | --- | --- |
|  | Positive | Negative |  |  |  |  |
| Positive | 43 | 7 |  |  |  |  |
| **Faeces** Negative | 0 | 20 | 1 | 0.86 | 0.741 | 1 |

The sensitivity test results of 100% indicates that all the positive results for SARS-CoV 2 that were identified via RT-PCR using saliva and faecal specimens were actually True positive. The Positive Predictive Value (PPV) was however 86% which shows that some True positive participants for SARS-CoV 2 were not able to be identified via the saliva sample.

The specificity result of 74.1% suggests that not all the participants who tested negative for SARS-CoV 2 using their saliva sample test were not having SARS-CoV 2 when tested using their faecal samples.

So, the False Negative were seven (7) participants representing 25.9%. However, the Negative Predictive Value (PPV) of 100% indicates that 100% of participants that tested negative using their saliva and faecal samples are truly negative for SARS-CoV 2.

### 4.6 Results of the Mean RT-PCR Cycle Threshold Values for

#### Nasopharyngeal/Oropharyngeal, Saliva and Faecal Samples

The Cycle Threshold (Ct) values of the RT-PCR test that was done on the pharyngeal samples of the fifty (50) SARS-CoV 2 positive participants ranged from 21.97 – 37.28 with a mean Ct value of 29.76. The saliva samples also presented with Ct values ranging from 20.71 – 37.94 with a mean Ct value of 29.82 whiles the faecal samples had Ct values ranging from 21.19 – 38.73 with a mean Ct value of 31.00.

**Figure 4:**
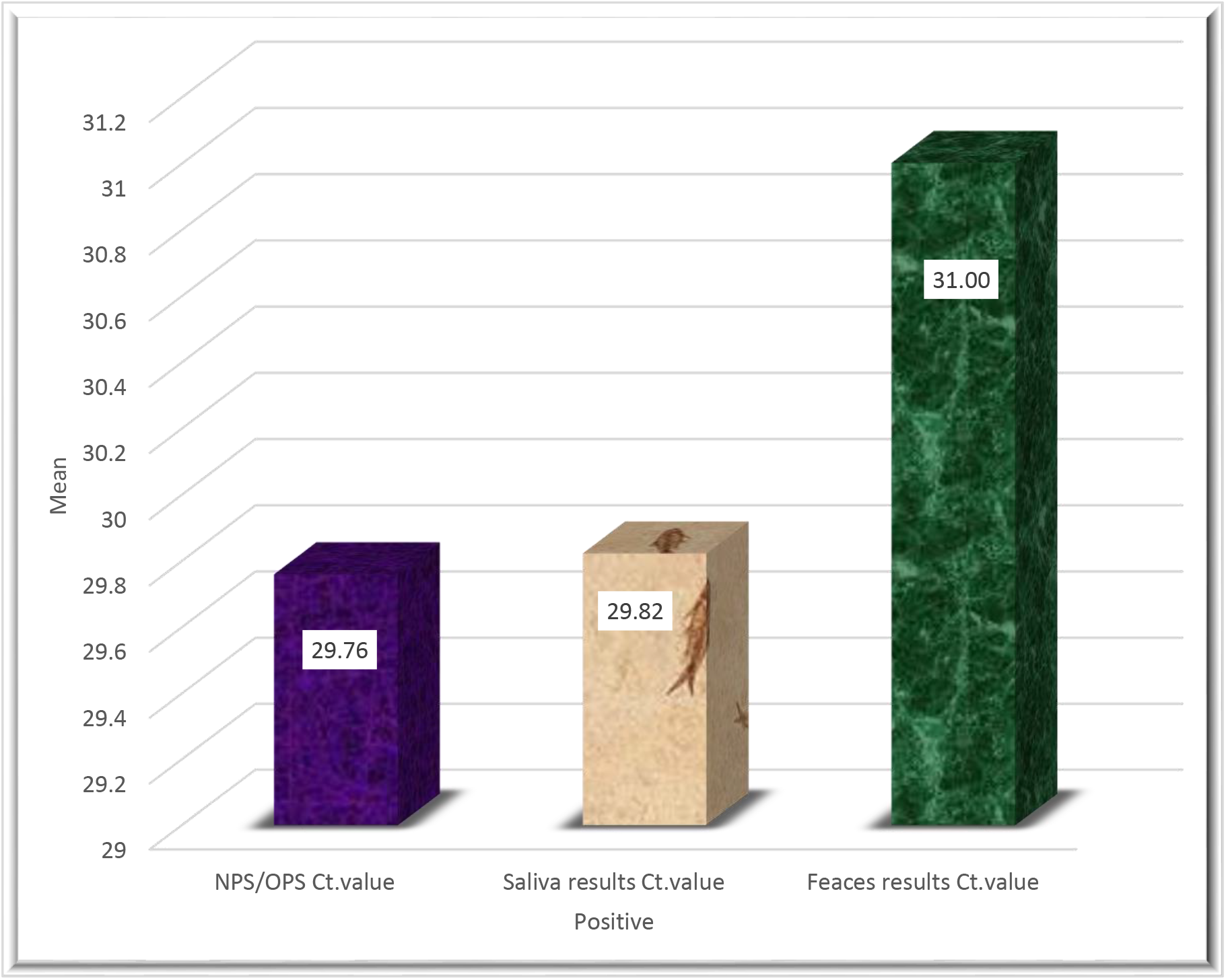
Bar Chart showing mean Cycle Threshold values for NPS/OPS, Saliva & Faeces.

## DISCUSSIONS

Looking at the results from this study in comparism with other studies carried out so far around the world by other researchers, it presents a very interesting scenario within the Ghanaian COVID-19 patient as well as other COVID-19 patients around the world that indeed, the possibility of an alternate sample and sampling technique to the current invasive, uncomfortable and risky pharyngeal sampling could be a reality one day. This is key especially for resource limited countries like Ghana and other sub-saharan countries as well who are looking at ways to reduce the cost associated with COVID-19 diagnostic testing as well as reducing nosocomial SARS-CoV 2 infection among Health workers who are at the forefront of the fight against this pandemic.

### 5.1 Viral shedding of SARS-CoV 2 in the Saliva and Faeces of Ghanaian COVID-19 Patients

This study has confirmed that indeed Ghanaian COVID-19 patients equally sheds SARS-CoV 2 in their saliva per the RT-PCR results obtained from the saliva samples taken from the study participants. This totally agrees with similar studies by Williams et al., (2020), Iwasaki et al., (2020), To et al., (2020), Hung et al., (2020) and Pasomsub et al., (2020) among other research works carried out by other Researchers but is in sharp contrast with the assertion made by Skolimowska et al., (2020) that saliva is an inferior sample especially when used in testing asymptomatic patients but rather confirms that saliva might indeed be a promising non-invasive alternative specimen for the diagnosis of 2019-nCoV infected patients.

The vital information this study further communicates is that, there might not be the need to expose suspected COVID-19 patients to the current invasive, uncomfortable and injury prone pharyngeal sampling that creates risky aerosols from the patients being sampled through coughing and sneezing after sampling.

A possible switch from pharyngeal sampling to saliva will also help in reducing drastically if not entirely, the number of Health workers getting exposed to SARS-CoV 2 through pharyngeal sampling with its resultant risky aerosols from the patients being sampled even when in full PPE. More importantly is the cycle threshold (Ct) values that saliva generated which was almost the same as the one generated by the pharyngeal swabs. This gave an indication that the possible viral load of SARS-CoV 2 in pharyngeal swabs is at par with the viral load in saliva samples further making a possible strong case for the use of saliva for testing.

This will also encourage more suspected patients to show up at our hospitals for testing because it is easy and more comfortable for a patient to give a small amount of his/her saliva than to be subjected to pharyngeal sampling.

Saliva as a sample may also be useful possibly in community surveillance testing as it is easier for someone to just give small amount of his/her saliva for testing than agreeing to be subjected to pharyngeal sampling. It is also comparatively cheaper to do saliva sampling than pharyngeal sampling because of the relatively cheaper cost associated with the consumables needed for saliva sampling than the cost associated with consumables needed for pharyngeal sampling technique. Furthermore, Governments and Policy makers not only in Ghana but around the world may save a lot of money that goes into the procurement of consumables that are used specifically for taking pharyngeal samples such as swab sticks, tongue depressors and viral transport medium.

Other cost cutting in caring for the Ghanaian public infected with COVID-19 could also be made from the number of eye goggles, N95 nose masks, coveralls and face shields as the quantities procured will reduce as some will no more be needed as PPE for pharyngeal sampling.

Alternatively, this study has equally confirmed that indeed Ghanaian COVID-19 patients not only sheds SARS-CoV 2 in their saliva but in their faecal matter as well per the RT-PCR results obtained from the faecal samples taken from the study participants. This totally agrees with similar studies by Xiao et al., (2020), Zhang et al., (2020), Lo et al., (2020), N. Zhang et al., (2020) and Jeong et al., (2020) among others. What this further means is that faeces could also possibly serve as an alternative sample for SARS-CoV 2 testing instead of the current invasive and discomforting pharyngeal sampling. Jeong et al., (2020) however came to an assertion that faeces have a good viral load which will enhance its diagnostic value for SARS-CoV 2 but this study differs from this assertion on that score based on the cycle threshold (Ct) values that the faecal samples generated which were on the high side indicating the possibility of a rather low viral load in the faecal samples as compared to saliva and pharyngeal samples.

It will also further help in reducing Health workers exposure to SARS-CoV 2 infection nosocomially through the aerosolization created by patients who are subjected to pharyngeal sampling via sneezing or coughing after sampling.

One interesting discovery this study made was that, the time the faecal samples were collected (morning, afternoon or evening) did not have any possible effect on the sample quality and viability. Just like other patients produce faecal samples for routine analysis, suspected COVID-19 patients can equally produce faecal samples by themselves for SARS-CoV 2 analysis.

It is cheaper compared to pharyngeal sampling because all that a suspected COVID-19 patient needs for sampling is a small stool container as used by other patients such as pregnant women undergoing routine stool examination as part of ante natal care.

The faecal viral shedding as evidenced by this study also supports the assertion reached by other researchers like Chen et al., (2020), that SARS-CoV 2 viral shedding in faecal samples of COVID-19 patients opens up the possibility of a feaco-oral transmission of the virus in areas where sanitation and human excreta disposal is a challenge.

### 5.2 Sensitivity and Specificity of using Saliva to test for SARS-CoV 2 as compared to Pharyngeal samples (Gold standard)

This study has also affirmed that indeed saliva is possibly as sensitive and specific for SARS-CoV 2 detection and it may be comparable to that of the pharyngeal swabs. This agrees with similar studies by Butler-Laporte et al., (2020), Pasomsub et al., (2020), Warsi et al, (2021), Vas et al, (2020) among other studies. What this mean is that, the possibility of getting false negative result from using saliva samples to do the testing instead of the pharyngeal samples is reduced. After all, no test procedure is absolute and this makes saliva a possible alternate sample in resource limited settings like Ghana. So, saliva can possibly be relied upon as a promising alternative sample for SARS-CoV 2 testing just like we currently rely on pharyngeal swabs or samples.

This further goes to possibly affirm the use of saliva for community screening for SARS-CoV 2 instead of the traditional invasive pharyngeal samples as people will be more willing to give a small portion of their saliva than to be subjected to pharyngeal sampling when they have no symptoms relating to SARS-CoV 2 infection. However, this finding was in sharp contrast with Skolimowska et al., (2020) who as part of his conclusion said that saliva may not be as sensitive a sample to use in SARS-CoV 2 testing in asymptomatic patients.

### 5.3 Sensitivity and Specificity of using Faeces to test for SARS-CoV 2 as compared to Pharyngeal samples (Gold standard)

The sensitivity and specificity of faecal samples as compared to pharyngeal samples obtained from this study goes to affirm that indeed, faecal samples could possibly be used in place of pharyngeal samples. This agreed with other studies carried out by Abdullah M et al, (2021) and Jeong et al., (2020) when it comes to the sensitivity and specificity of using faeces to test for the presence of SARS-CoV 2.

The possibility of getting a false negative result is reduced if not completely erased. So faecal samples could also be a promising alternative sample to the pharyngeal swabs or samples because of its very good sensitivity and specificity.

Although the mean cycle threshold (Ct) value for the faecal samples was high as compared to the pharyngeal samples giving an indication of a lower viral load with less chances of detection, the RT-PCR was still able to detect SARS-CoV 2 within the faecal samples which goes to affirm that, it may be a sample that can possibly be relied upon for SARS-CoV 2 testing in resource limited settings like Ghana.

## CONCLUSIONS AND RECOMMENDATIONS

### 6.1 Conclusion

In conclusion, this has been a good experimental case study and the first of its kind in Ghana so far as the search for an alternative sample for SARS-CoV 2 among Ghanaian COVID-19 patients is concerned. It has really lived up to its objectives in providing some preliminary data to support further research into the possible use of saliva or faeces as an alternative sample for SARS-CoV 2 testing among Ghanaians and other patients around the world within the current pandemic period instead of the current pharyngeal sampling which comes with a much higher cost and risk of exposure to SARS-CoV 2 infection among Health workers doing the sampling especially. It also comes with much comfort to the patients providing the sample just like they have been providing their faecal or other samples themselves for routine and specialized laboratory diagnostic tests over the years. All the possible injury to the patients during the pharyngeal sampling, the sneezing, coughing or even fear of pain will all become a thing of the past.

Self-collected saliva or faeces carry no risk to the Health worker in terms of their exposure to SARS-CoV 2 from a suspected patient during the sampling process. This is because no aerosols are produced from self-collected saliva or faeces. Even with weak, severely sick and frail patients that may need some assistance from Health workers to take the saliva or stool sample, the risk of exposure is much less than if pharyngeal sampling process was used. Simple sterile plastic stool containers can be used for both the saliva and faecal collection in resource limited settings like Ghana. This will even have an impact on the consumables’ component of the cost build up per one SARS-CoV 2 RT-PCR test being currently charged by the testing centers in Ghana.

### 6.2 Recommendation

This study recommends that Government of Ghana, our Development partners and the scientific community pays some attention to ongoing research on COVID-19 especially those researches that aim at finding an alternative sample for testing for the virus aside the current pharyngeal samples as this will go a long way to at least reduce the spread of the virus among Health workers who are most at risk of contracting SARS-CoV 2 with our current pharyngeal sampling process.

It will also encourage more patients experiencing mild to moderate symptoms to show up at Health facilities for testing instead of staying at home due to the fear of pain and discomfort associated with the pharyngeal sampling process.

It is further recommended that this study be expanded in terms of the sample size and replicated at the different testing sites in Ghana, so as a Nation, we can come out emphatically to say if indeed saliva or faecal sample can be an equally good surrogate sample for testing SARS-CoV 2 aside the pharyngeal samples in Ghana. This when done will have the potential of effecting a policy change in the sample of choice or samples a suspected patient can choose from when asked to produce a sample for SARS-CoV 2 testing in Ghana.

Early morning saliva before oral health is performed by a patient and saliva taken after oral health or any other time after oral health could also be explored further as part of our efforts to find the best saliva sampling time and sampling condition for the best sample quality and reliability should saliva be accepted as the best alternative sample as against faeces in SARS-CoV 2 testing. We should also be looking at culturing the virus which is shed in saliva and stool/faecal samples as a country to find out if the virus being shed is a live infective SARS-CoV 2 as was done in other studies. It is also recommended that as a Nation, we explore further the possibility of a feaco-oral transmission of SARS-CoV 2 among our populace by picking samples from our sewage system and checking if they will test positive for SARS-CoV 2 as well as if the SARS-CoV 2 the faecal samples might contain is a live infective one.

## Data Availability

All data on this study is readily available for sharing on request

https://www.researchgate.com

## DATA AVAILABILITY

Data concerning this study will be made available to the scientific and research community upon request to the principal investigator.

## ACKNOWLEDGEMENT

This study would like to acknowledge the immense support of Dr. Lydia Sarponmaa Asante, Dr. Albert Dompreh and Dr. Laud Anthony Basing for their time, patience, dedication and the direction they gave to make this study a success. The entire staff of the Infectious Disease & Research Laboratory (IDRL) are hereby acknowledged especially their Acting Technical Head, Mr. Kwabena Adjei Asante, Miss Sylvia Karikari and Mr. Albert Adubofour for their immense support.

This study would also like to acknowledge Dr.Chris Oppong of the Emergency Medicine Directorate/IDHA and Mad. Faustina Acheampong, Nurse in Charge of HIIU for their contribution during the sampling for patients within their various Units. Acknowledgement also goes to the Laboratory Services Directorate and its entire management team led by Dr. Ernest Adjei and the entire staff of the Clinical Parasitology Unit-KATH for their immense support in making this study a success.

## STUDY TEAM

Ernest Badu-Boateng (PI), Lydia Sarponmaa Asante (PhD), Albert Dompreh (PhD), Laud Anthony Basing (PhD), Kwabena Adjei Asante, Sylvia Karikari, Chris Oppong (MD), Albert Adubofour, Faustina Acheampong (CNO)

